# DETERMINANTS OF HUMAN IMMUNODEFICIENCY VIRUS (HIV) TRANSMISSION AMONG CHILDREN BORN TO HIV-INFECTED MOTHERS ON PMTCT PROGRAM IN KILIFI COUNTY: A CASE CONTROL STUDY

**DOI:** 10.1101/2023.06.12.23291301

**Authors:** Rukiya Mohammed Koriow, Julius Otieno Oyugi, Christine Muasya, Rodgers Onsomu Moindi

**Affiliations:** University of Nairobi Institute of Tropical and Infectious Diseases, Nairobi, Kenya; Department of Medical Microbiology, College of Health Sciences, University of Nairobi, Director University of Nairobi Tropical and Infectious Diseases (UNITID) Nairobi, Kenya; University of Nairobi, Department of Clinical Medicine and Therapeutics Nairobi, Kenya; Research unit, Department of Health, County Government of Kilifi

**Keywords:** Kilifi, HIV positive mothers, PMTCT, ANC, Infant HIV Status

## Abstract

**Background:** Elimination and prevention of mother to child human immunodeficiency virus (HIV) transmission remains a global public health concern. Identification of significant risk factors of mother to child transmission is essential in reducing the burden of pediatric HIV in terms of cost of treatment, morbidity and mortality. Only a few studies have been done to demonstrate the importance of Prevention of Mother to Child Transmission (PMTCT) cascade, including the indicators and how missing one of the crucial steps of the cascade could increase the risk of infant HIV transmission. This study seeks to determine maternal risk factors related to HIV transmission among children born to HIV-infected mothers enrolled in PMTCT program in Kilifi County between July 2018 and June 2019.

**Methods:** This was a retrospective unmatched case control study. Systematic random sampling was used to select medical records of 183 exposed infants (61 cases and 122 controls). A structured tool was used to collect data. Proportions and frequency distribution was done in univariate analysis and in bivariate analysis, chi-square and logistic regression (odds ratio) was used to determine any association and strength of association between categorical variables.

**Results:** Receiving maternal HAART in ANC was established as the major determinant of a favorable HEI outcome in this study. Infants whose mothers had received maternal HAART in ANC were 52.55 (14.89 - 185.48) times more likely to be HIV negative compared to those whose mothers did not receive maternal HAART in ANC. Disclosure of HIV status by the mother at ANC was 3.18 (1.28 - 7.90) times more likely to determine a favorable outcome of the HEI compared to non-disclosure. When adjusted for other factors, it was 6.27 (1.20 - 20.05) times more likely to determine a favorable outcome.

**Conclusion:** The study established the importance of PMTCT service delivery in reduction of vertical transmission of HIV. Women who defaulted on the PMTCT cascade services had higher odds of having a HIV positive infant compared to women who accessed all the PMTCT services at the recommended time.

## Introduction

### Background

Among all age groups, Human Immunodeficiency Virus/Acquired Immune Deficiency Syndrome (HIV/AIDS) is a lifelong disease with significant morbidity and mortality. Globally, over 38 million people in 2018 were living with HIV/AIDS, with two-thirds of this population and 2.1 million children below 15 years residing in Sub-Saharan Africa (1). Approximately 75 million people worldwide have become infected with 32 million people having been reported to die due to AIDS related illnesses since the beginning of the epidemic. Out of these, 770,000 died in 2018. However, the number of deaths has significantly reduced since 2004 from the peak of 1.7 million to 1.4 million reported in 2010 (2). New HIV infections among children less than 15 years old have globally reduced (47%) from 280,000 in 2010 to 160,000 in 2018, which reflects a 42.9% reduction (3); (4). Approximately 1% of pregnant women worldwide are HIV infected. In 2015, 1.4 million women of reproductive age living with HIV became pregnant and majority were from sub-Saharan Africa (5). HIV transmission from an infected mother to her child i.e. vertical transmission may occur during pregnancy, childbirth or while breastfeeding (6). More than 90% of the mother to child HIV transmission cases are due to vertical transmission (7). This transmission can entirely be eliminated. However, this has proved to be a challenge, especially in some of the 22 priority countries listed by the Global Plan to accelerate efforts in prevention of transmission (PMTCT) of HIV and syphilis among the newborns from the mother (8)(9). In 2015, six African countries (Botswana, Mozambique, Namibia, South Africa, Swaziland and Uganda) listed among the priority countries met the 90% target of reducing mother to child transmission. Cuba, in mid-2015, was the first country to eliminate MTCT. Similar achievements were noted in the Asia and pacific region of Thailand in 2016, with Belarus and Armenia reporting less than 5% transmission rate (10). Kenya is among the sub-Saharan Africa countries most affected by HIV/AIDS pandemic, with at least 2 million adults living with HIV/AIDS. In 2017, it was reported that there were 110,000 children living with HIV and 8,000 new infections, with a reduction from 25,000 to 13,000 in 2005 and 2010 respectively. In addition, 54,000 HIV positive mothers were on PMTCT program and 76% had access to antiretroviral treatment (ART), which was significantly higher compared to 2010 (56%) (5). Much has been done in elimination of mother to child transmission (e-MTCT) reducing new infections and increasing access to ART among pregnant women. This is due to adoption and implementation of United Nations four-pronged strategy for PMTCT (4); (12). Kenya, which is among the 22 priority countries listed, has made tremendous efforts, which have seen the mother to child transmission rate of HIV infection reduce from 16% in 2013 to 8.3% in 2015 (13). However, this success was short-lived as the MTCT rate increased to 11.5% by 2017 and up to 12.4% by 2018 (12). Kenya adopted the Global Plan and formulated the Kenya Framework for EMTCT transmission 2016-2021 to give the country strategic guidance on validation of goals for elimination of mother to child transmission by 2021 (14). To realize this dream, Kenya has to achieve more than 95% on each of the process indicators depicted on the PMTCT cascade (12). Previous studies conducted in sub-Saharan African countries including Kenya mainly investigate the possible determinants of vertical transmission of HIV infection among HIV positive mothers, only a few demonstrate the importance of the PMTCT cascade, including the indicators, and how missing one of the crucial steps of the cascade could increase the risk of HIV transmission from the mother to the infant (15). Kilifi, a predominantly rural county in the coastal region of Kenya has a HIV prevalence of 4.4%. It is one of the counties identified in the Kenya framework for elimination of mother to child transmission (2016-2021) due to the upward increase in MTCT rate from 7.7% (2015) to 8.3% (2018) (16); (14). During the financial year 2018-2019, Kilifi County had a total of 62 HIV-exposed infants who were confirmed to be HIV infected despite many efforts geared to reverse the incidence of pediatric HIV. Due to this, the county has set up several innovative ways to improve key PMTCT indicators and despite laudable progress, the county’s goal to eliminate HIV remains unaccomplished. The worrying numbers show a default in the PMTCT cascade by the HIV positive women.

This study, aimed to determine the risk factors associated with the outcomes of HIV exposed infants with the uptake of PMTCT indicators among the HIV-positive mothers based on the hypothesis that HIV exposed infants who become HIV infected are born to HIV positive women who default on the PMTCT cascade.

### Study goal

To identify determinants of mother to child transmission among children born to HIV positive mothers on PMTCT Program in Kilifi County between July 2018 and June 2019

### Specific objectives

1. To determine the association between ANC attendance of mothers and outcome of their HIV exposed children.
2. To determine the association between HIV testing of mothers at ANC and outcome of their HIV exposed children.
3. To determine the association between Maternal HAART and Infant prophylaxis and outcome of their HIV exposed children
4. To determine the association between skilled delivery of HIV positive mother and the status of her child.

### Conceptual framework

**Figure 1:**
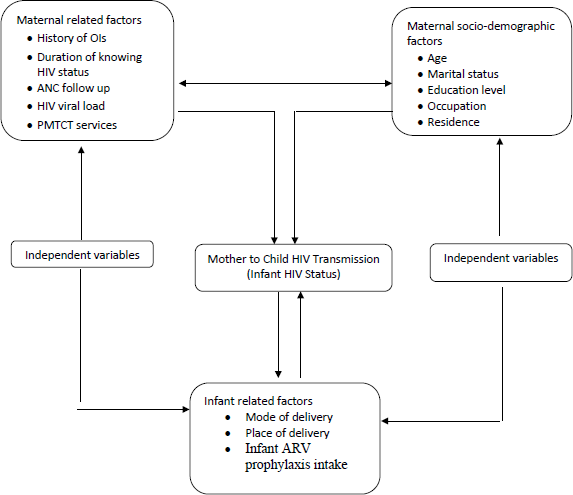
Diagrammatic representation of the conceptual framework

### Methodology

#### Study design and period

This was a retrospective unmatched case control study, based on patient records. The study participants did not necessarily have the same characteristics, but were from the same population. Selection of the study participants was based on HIV positive mothers who were enrolled in the PMTCT program and their index children. The mothers had a confirmatory HIV test done between July 2018 and June 2019 in Kilifi County.

A Case was defined as a HIV positive mother who was enrolled in the PMTCT program with her child who was tested and confirmed to be HIV infected between July 2018-June 2019 while a control was defined as a HIV positive mother who was enrolled in the PMTCT program with her child who was tested and confirmed to be HIV negative between July 2018-June 2019. The study period was between May 2020 to December 2021.

To control bias; the controls were randomly selected during the study period as the cases, hospital records were used and also during staistical analysis.

#### Study site and population

The study was carried out in Kilifi County which is located at the coastal region of Kenya. The county has a total population of 1,545,211 (17). It is divided into 7 sub-counties with a total of 143 public health facilities and 151 private health facilities. Out of these, 173 offer PMTCT services to approximately 7% of the HIV infected female population. The study population included all HIV positive mothers whose HIV exposed child or children had a confirmatory HIV test result, positive for cases and negative for controls between July 2018-June 2019 in the different healthcare facilties.

### Variables

#### Dependent Variables

Infant HIV status confirmed by DNA PCR testing (mother to child HIV transmission).

#### Independent Variables

ANC attendance of mother, Knowledge of HIV status of mother, Maternal HAART and Infant prophylaxis and place of delivery of mother.

### Inclusion criteria

- A HIV positive woman enrolled in the PMTCT program in Kilifi County
- A HIV positive mother whose child had a confirmatory HIV test, positive for cases and negative for controls between July 2018 and June 2019.

### Sample size

To determine the sample size, Epi Info Version 3.01 Stat calc function of sample size calculation for unmatched case-control study was used (18). At 95% confidence interval and 80% power estimates were derived from studies done on determinants of mother to child HIV transmission in Ethiopia (19). Least odds ratio of 1.80 assuming 32.4% and 74.9% of mothers in the cases and control group respectively exposed to PMTCT follow up at least four times. Considering a 1:2 ratio of cases to controls, a sample size of 183 mother baby pairs (61 cases and 122 controls) was the calculated sample size.

### Sampling technique

Stratified random sampling method was applied to select study subjects from the different county healthcare proportionately to their study population size. Cases and controls were selected by accessing the available digital/online medical registry. In reference to infant DNA HIV results, strata (the different healthcare facilities within the county) numbers were identified from the county population and by simple random sampling; cases and controls were selected proportionately (Fig. 2).

**Fig. 2.**
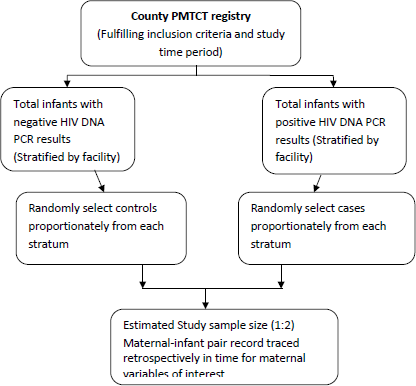
Selection of cases and controls in Kilifi County

### Data collection

To collect data, a researcher structured tool consisting of socio-demographic, infant and maternal characteristics was used. Data was extracted from ANC, Maternal and Postnatal registers. Data was collected by clinical nurses who had been trained and had at least 1 year experience in PMTCT service delivery at the selected facilities. The data was shared in MS Excel in a password controlled computer and backed up in a flash disk owned by the principal investigator.

Clients individual information including names, file and clinic number were excluded when the data collection tool was recorded. The authors did not have information that could identify individual participants during or after the study.

### Data management

Data in MS Excel was cleaned and assessed for completeness and analysis done using MS Excel and IBM SPSS Statistics version 21. For categorical variables such as study categories (cases and controls) and gender, frequency and proportions was used. Chi-square test and logistic regression was done to assess any relationship between the dependent variable and categorical independent variables such as the breast infection, mixed feeding ANC visits. Odds ratio (OR) was used to estimate the strength of association between the exposure variables like ANC follow up, duration of knowing HIV status and mother to child HIV transmission. Data is presented in tables.

### Ethical consideration

Ethical review of the proposal was done by KNH-UON Ethics and Research Committee. This was a retrospective study based on secondary data; therefore, a waiver for informed consent was requested. Once the study was approved (P374/07/2020); permission and approval to conduct the study was also sought from the Kilifi County Health Management Team, Medical Superintendent and the Medical Records Officer in Charge, Kilifi County. Patient information including names, file and clinic number were excluded and the data collection tool recoded. In addition, the patient’s medical records will be maintained in a safe lockable cabinet and destroyed after 5 years by burning to maintain confidentiality. Patients may not benefit directly from the study but the report will be shared with clinicians and relevant stakeholders to improve general patient care and management. The probable risk associated with the study includes loss of patient information, although the risk will be mitigated by use of trained healthcare workers and use of password controlled computers and lockable cabinets.

## Results

### Demographic Characteristics

#### Gender and Age distribution

The study sampled 161 infants enrolled in HIV Exposed Infant (HEI) program across various health facilities within Kilifi County. Of these, 49% were male and 51% females. Across the infant HIV test outcome categories, there were nearly same proportion of males as females though only 59 participants were sampled as Positive and 101 as negatives.

In terms of age in weeks at enrolment into the HEI program, 56% of the infants were between 4 and 24 weeks while 33% of the infants were less than a month old. Of all the HIV positive infants 58% were between 4 and 24 weeks of age while 17% were less than 4 weeks old. Of all the HIV negative infants 55% were between 4 and 24 weeks old while 43% were less than a month old at time of enrollment.

The table below summarizes the social demographic characteristics.

**Table 1:** Summary of demographic characteristics of infants sampled

| Sex: | Infant HIV Test Outcome: |  |  |  | Total<br>(n=160) |  |
| --- | --- | --- | --- | --- | --- | --- |
|  | Positive<br>(n=59) |  | Negative<br>(n=101) |  |  |  |
| Male | 29 | 49% | 50 | 50% | 79 | 49% |
| Female | 30 | 51% | 51 | 50% | 81 | 51% |

| Age of infant at Enrolment into HEI<br>Program (weeks) | Infant HIV Test Outcome: |  |  |  | Total<br>(n=161) |  |
| --- | --- | --- | --- | --- | --- | --- |
|  | Positive<br>(n=60) |  | Negative<br>(n=101) |  |  |  |
| <4 | 10 | 17% | 43 | 43% | 53 | 33% |
| 4-24 | 35 | 58% | 55 | 55% | 90 | 56% |
| 25-36 | 8 | 13% | 1 | 1% | 9 | 6% |
| 37-72 | 7 | 12% | 0 | 0% | 7 | 4% |

#### Other Infant Variables

- Identification point of care
- PCR test results
- Infant prophylaxis regimen

In terms of identification, 81% of the infants were identified through MCH/PMTCT program, 7% were identified through OPD and the others were identified through CCC, Maternity and in Pediatrics ward. Among all the positive infants, 58% were identified through MCH/PMTCT and 17% through OPD, 10% through pediatrics ward and 7% in CCC. Among those confirmed to be negative, 95% of them were identified through MCH/PMTCT services.

Majority (90%) of the infants enrolled received their PCR test results within the 1^st^ 6 weeks or at first contact. Only 10% of the infants were not provided PCR test within the initial 6 weeks or at first contact overall. Among those who tested positive, 20% did not receive their PCR test in the initial 6 weeks or at first contact. This proportion stood at 4% for those confirmed to be negative

Out of all the infants enrolled only 4 did not receive infant prophylaxis and cotrimoxazole. Of those that had received infant prophylaxis, 88% had received both NVP and AZT while 12% received only NVP. 91% of the positive infants were on NVP and AZT while 9% were on NVP alone. 87% of the negative infants were on NVP and AZT while 13% were on NVP alone.

Table 2 below summarizes other infant variables

**Table 2:**
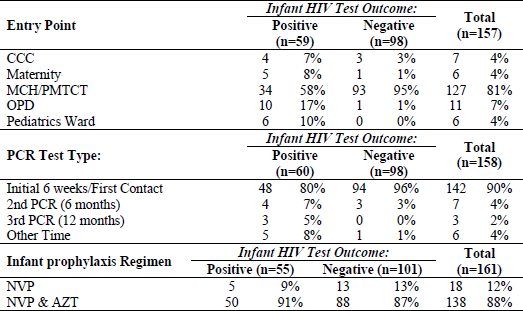
Other Infant Variables

#### Maternal Variables

- ANC attendance
- Knowledge of HIV status
- Maternal HAART
- Disclosure of Maternal HIV Status
- Place of delivery (Skilled delivery or home delivery)

Majority (88%) of mothers to all infants sampled in the study had attended ANC at least once. All the mothers who had a HIV negative infant had attended ANC while 33% of those who had a HIV positive infant did not attend ANC.

Most of the mothers (95%) of the infants sampled knew their HIV status prior to attending ANC. This proportion was the same in both cases and control group.

On disclosure of HIV status, mothers to 86% of all the infants confirmed disclosing their HIV status. Among the HIV positive infants, 76% had disclosed while for the HIV negative infants, 91% had disclosed.

Overall, most mothers (80%) had disclosed their status to their partners. More specifically, this was 88% among the mothers to 88% of the infants that were positive and mothers to 76% of the infants that were negative from the HEI outcomes.

Seventy-five (75%) of the mothers of all infants sampled received maternal HAART. Majority (62%) of the mothers whose infants tested positive for HIV did not receive maternal HAART while this proportion was only 3% among the mothers of HIV negative infants.

For the mothers that had started maternal HAART in ANC, 47% started at 1^st^ trimester. Among positive, the proportion of those initiated on HAART within 1^st^ trimester was 23% compared to 53% of those in the negative arm started on HAART around the same reference period.

Of these deliveries, 82% of the mothers to all the sampled infants delivered at the health facility compared to 18% at home. Among the positive, home deliveries were 42% compared to 3% among negatives while hospital deliveries were 58% among the positive compared to 97% among negatives.

**Table 3:**
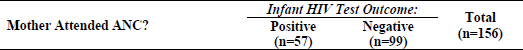
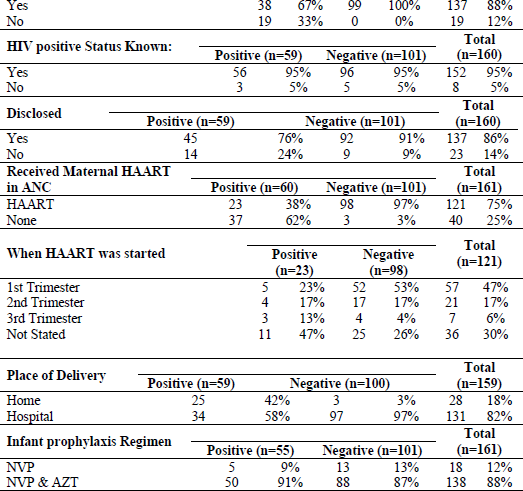
Summary of Maternal Variables

### Bivariate and Multivariate Analysis

The study established a significant relationship between the HIV status of the HIV exposed infant and the ANC attendance of the mother (p=0.00) and the mother receiving HAART in ANC (p=0.00) using the chi-square test at a 5% level of significance. Only receipt of maternal HAART in ANC was established as the major determinant of a favorable HEI outcome in this study. On the nature of the relationships between the outcome of the HEI and the independent factors, disclosure of HIV status by the mother at ANC was 3.18 (1.28 - 7.90) times more likely to determine a favorable outcome of the HEI compared to non-disclosure at a 5% level of significance when taken independently. When adjusted for other factors, it was 6.27 (1.20 - 20.05) times more likely to determine a favorable outcome of the HEI at 1% level of significance. On maternal HAART at ANC, infants whose mothers had received maternal HAART in ANC were 52.55 (14.89 - 185.48) times more likely to be HIV negative compared to those whose mothers did not receive maternal HAART in ANC at 1% level of significance and taken independently of other factors. This came down to 30.23 (7.56 - 120.96) times when adjusted for other factors at the same level of significance.

**Table 4:**
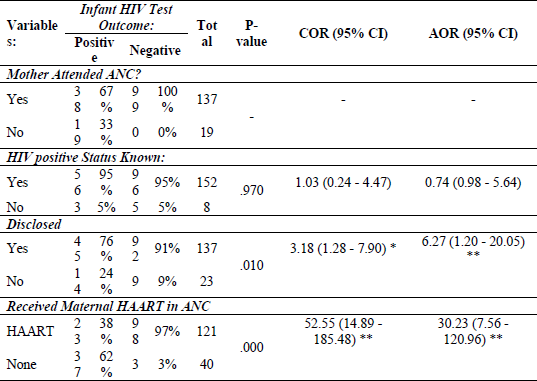
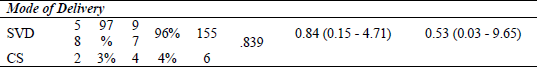
Summary of Bivariate and Multivariate analysis

## Discussion

ANC services provides an opportunity for key interventions in prevention of vertical transmission of HIV. Such interventions include early diagnosis of the HIV positive pregnant woman, prompt initiation of HAART, Viral load monitoring and Enhanced Adherence Counselling when necessary, psychosocial support and health education on importance of skilled delivery. All these interventions contribute towards a HIV negative infant.

The current National ANC attendance is at 74%, there is need to improve this to 95% as ANC provides an opportunity for HIV testing and initiation of HAART for the HIV positive mother. Importance of improving this indicator is evidenced by Kenya National Bureau of Statistics/Government of Kenya, 2015 which showed that only 19% of Kenyan women visit the ANC within the first 4 months of pregnancy and 31% make their initial visits at 4-5months and 6-7 months respectively and only 68% visit the clinic at least 4 times.

On attendance of antenatal care clinics, the study found that all mothers who had HIV negative infant had attended ANC at least once while for the mothers who had a HIV infected infant only 67% had attended at least 1 ANC visit. This finding demonstrates the importance of Antenatal care clinics and is consistent with literature on access to ANC services and reduction of mother to child transmission rate. In a study done by *Gunn et al* on Antenatal care and uptake of HIV testing among pregnant women in Sub Saharan Africa, it was established that women who had access to ANC services accessed HIV testing services which is the entry point in PMTCT care cascade (6). In another study done in Brazil, it showed that lack of antenatal care and late diagnosis underlie substantial gaps in PMTCT cascade (20) No studies were found that do not agree with importance of ANC in prevention of vertical transmission of HIV.

Our study evaluated the relationship between knowledge of the mother’s HIV status and outcome of their infants. The study sought to find out whether women who know their HIV positive status early in pregnancy had lower risk of having a HIV positive infant on the basis that interventions of PMTCT would be started early. Identification coverage of HIV positive women in reproductive age is important in reducing MTCT of HIV. Women who know their HIV status are more likely to have started HAART and are more likely to have achieved viral load suppression at the time of conception, during pregnancy and breastfeeding. Planned pregnancies among the HIV positive women decreases the odds of a mother transmitting HIV virus to her baby. The study did not show any difference in knowledge of HIV status of the mother and outcome of their children as 95% of women in both the control and cases group knew of their status before ANC. This finding was similar to a study done in Western Kenya which aimed at describing the mother baby pairs enrolling in PMTCT program. It showed that 68% of Known Positive women enrolled in PMTCT were not on ART at time of conception despite their knowledge of their HIV status. For those on HAART, a significant number (14%) had an unsuppressed viral load. All which suggested that knowledge of HIV status alone may not contribute to prevention of mother to child transmission (21) This finding was however contrary to a study done in South Africa that showed that women aware of their HIV status before the index pregnancy were more likely to receive PMTCT and to test their infants early hence have a HIV negative infant. (22) Several case control studies looking at determinants of mother to child transmission showed a positive correlation between women who did not know their status and mother to child transmission of HIV infection (23) (20)

Disclosure of one’s HIV status improves uptake and retention in prevention of mother to child transmission of HIV services. Disclosure of HIV status allows the mother to have psychosocial support this in turns leads to increased adherence to PMTCT services. Our study evaluated whether disclosure of the mother’s HIV positive status had any relationship to the outcome of the HIV status of the infant. On disclosure of HIV status, mothers to 86% of all the infants confirmed disclosing their HIV status. Among the HIV positive infants, 76% had disclosed while for the HIV negative infants, 91% had disclosed their status. This finding is consistent with a study done in Kenya that assessed HIV positive status disclosure and use of essential PMTCT and Maternal Health services. It established that women living with HIV who had not disclosed to anyone had the lowest levels of PMTCT service utilization (24) In another study done in Kenya looking at the prevalence and correlates of non-disclosure of maternal HIV status to male partners; it established that non-disclosure was associated with lower use of PMTCT services and facilitating maternal disclosure to male partners may enhance PMTCT uptake (25).

Prompt HAART initiation is associated with rapid Viral load suppression consequently reduction in vertical transmission of HIV. Our study evaluated the relationship between uptake of maternal HAART among the mothers and HIV status outcome of their infants. It also looked at the relationship between uptake of infant prophylaxis among HIV exposed infants and their HIV status. The study found that 75% of all the mothers were on HAART while 25% of them were not on HAART. On relationship between maternal HAART and HIV status of the HIV exposed infant it was established that 97% of the mothers of the infants in the control group were on HAART while only 38% of the mothers of the infants in the cases group were on HAART. 62% of the infants who became HIV infected were born of mothers who were not on HAART. Several studies have shown the importance of maternal HAART in prevention of vertical transmission. As lack of Maternal HAART leads to maternal HIV viremia which increases the odds of vertical transmission (21). In another study done in Nigeria looking at the impact of Maternal HAART in its prevention of vertical transmission of HIV showed that, none of the exposed children born to HIV positive mothers on HAART were vertically infected with HIV-1 and HIV-2 (26).

Early initiation of maternal HAART by HIV positive pregnant women has been shown to reduce viral load of HIV to undetectable levels and hence reduce vertical transmission of HIV. Our study also agreed with this as 53% of the infants who confirmed to be HIV negative were born of mothers who were initiated on HAART during 1^st^ trimester compared to the 23% of HIV positive infants whose mothers initiated HAART in 1^st^ trimester.

On adherence to maternal HAART, 71% of the mothers were rated to have good adherence while 16% were rated poor. 94% of the infants who tested and confirmed to be negative were born of mothers who were rated to have good adherence. A study done in neighboring Ethiopia established that poor adherence to maternal HAART was associated with an increased risk of vertical transmission of HIV(23)

Skilled delivery is defined as delivery conducted by trained healthcare personnel. Labor and delivery poses the greatest transmission risk of HIV with 10-20% exposed infants being infected during this time. The study evaluated the relationship between skilled delivery and outcome of the HIV exposed infant. The study established 82% of the mothers delivered at the health facility compared to 18% at home. Among the positive, home deliveries were 42% compared to 3% among negatives while hospital deliveries were 58% among the positive compared to 97% among negatives. Similar studies done established the importance of skilled delivery in prevention of mother to child transmission of HIV (21) (23) (27).

## Conclusion

Similar to literature reviewed from other countries this study established the importance of PMTCT service delivery in reduction of vertical transmission of HIV. ANC attendance, early diagnosis of maternal HIV, prompt initiation and adherence to HAART, skilled delivery of the HIV positive pregnant woman were all associated with reduction of mother to child transmission rate. Based on the findings it was established that women who defaulted on the PMTCT cascade services had higher odds of having a HIV positive infant compared to women who accessed all the PMTCT services at the recommended time. This agreed with a study by *Hamilton et al* PMTCT cascade analysis which showed that new HIV infections among children were seen among women who did not complete their cascade (28).

Other variables that showed association were male partner involvement and involvement of the mother in psychosocial support groups. These findings are similar to studies which looked at such determinants (23) (20) (21) (28).

### Recommendation

There is need to emphasize the importance of attending ANC clinic for all pregnant women.

There is need to emphasize the importance of all the steps of the PMTCT Cascade to HIV positive mothers attending ANC.

There is need to ensure that all women who test positive for HIV are started on HAART and followed up for adherence.

### What is already known about this topic

- Vertical transmission of HIV can be eliminated and ANC services provides an opportunity for key interventions in prevention of vertical transmission of HIV.
- Women who know their HIV status are more likely to have started HAART and are more likely to have achieved viral load suppression at the time of conception, during pregnancy and breastfeeding. Planned pregnancies among the HIV positive women decreases the odds of a mother transmitting HIV virus to her baby.
- Prompt HAART initiation is associated with rapid Viral load suppression consequently reduction in vertical transmission of HIV.

### What this study adds

- The findings from this study provide an insight on the importance antenatal clinic attendance in diagnosis of HIV and on initiation of HAART among pregnant women thus preventing vertical transmission of HIV
- The findings from this study highlights missed opportunities and its consequences in Prevention of Mother to Child transmission of HIV.

## Data Availability

All relevant data are within the manuscript and its Supporting Information files.

## Acknowledgements

We deeply appreciate Fauz Ibrahim and his team for their assistance in data collection and their commitment in this study.

